# Disease prevalence, health-related and socio-demographic factors in the GCAT cohort. A comparison with the general population of Catalonia

**DOI:** 10.1101/2023.09.08.23295239

**Authors:** Natalia Blay, Lucía A Carrasco-Ribelles, Xavier Farré, Susana Iraola-Guzmán, Marc Danés-Castells, Concepción Violán, Rafael de Cid

## Abstract

**Background:** Population-based cohorts play a key role in epidemiological studies. However, it is known that volunteer cohorts include a healthy volunteer bias. Assessment and characterization of this bias is needed to extrapolate results to the general population. Here, we assess the bias of the population-based cohort GCAT, encompassing 20 000 adult participants from Catalonia with electronic health record data. The aim of this study is to compare the GCAT cohort with its age-matching Catalan population, to assess their representativeness, as well as determining the weights to make results generalisable.

**Methods:** Statistical comparisons until 2019 in multiple variables across sociodemographic, lifestyle, diseases and medication domains were performed by stratified analysis with Fisher’s exact test and t-test. Electronic health records of Catalonia (SIDIAP), and registers from the statistics institute of Catalonia (IDESCAT) and Spain (INE) were used to make the comparisons. We generated weights accounting for sociodemographic, lifestyle and multimorbidity factors.

**Results:** GCAT cohort is enriched in women and younger individuals, with higher socioeconomic status, more health conscious and healthier in terms of mortality and chronic disease prevalence. We have shown that this bias can be corrected with weighting techniques, providing a more representative sample of the general population.

**Conclusions:** The application of multidomain weights, encompassing not only sociodemographic aspects, but also lifestyle and health-related variables, has effectively diminished the observed bias in disease prevalence estimates within the GCAT cohort. This correction has led to an enhancement of the cohort’s representativeness, rendering it more akin to the general population of Catalonia.

## Introduction

Complex diseases are caused by a combination of multiple genetic factors, environmental exposures and lifestyle habits, and they represent the majority of diseases [1, 2]. Some of these risk factors have not yet been identified due to its complex interplay, which increases with multimorbidity, the co-occurrence of more than one complex disease in the same individual [3]. To this purpose, longitudinal population-based cohorts play an important role, as they usually involve a large number of well characterised individuals, representative of the population. These characteristics are not only important for risk factors detection, but to develop general prevention strategies and treatments [1].

GCAT cohort is an adult population-based cohort from Catalonia, at the north-east of Spain. It comprises almost 20 000 volunteer individuals between 40-65 at the recruitment time (2014-2018). At that time, they completed an extensive questionnaire about lifestyle habits and diseases, provided blood samples, and their anthropometric measures were determined. In addition, they provided informed consent to access their electronic health record (EHR) data and geocoded residences for environmental exposure, that, together with yearly follow-up questionnaires, allows a deeply phenotyping of the cohort and the generation of multiomics data, including genotypes, Whole Genome Sequencing (WGS) data, metabolome, proteome and epigenotypes for a number of individuals of the cohort [4].

EHR-linked population-based cohorts offer an extremely rich longitudinal characterization of individual-level health factors, which can help determine the individual relative risk for certain complex diseases. This is key for precision medicine, helping with the prevention of complex diseases in high-risk patients, and personalized treatment after the diagnosis [2, 5]. Nonetheless, population-based cohorts commonly exhibit biases due to the healthy volunteer effect, which is a form of selection bias wherein volunteer participants tend to possess better health compared to the general population [6, 7]. This can challenge the generalisation of the results obtained from this type of cohorts, as there are some groups of population that are not well represented in the cohort, such as low socioeconomic level or poor-health individuals. This type of bias can be avoided using weighting approaches, such as raked-weights [8], which allow the generalisation of results to the entire population.

In this study, we compare the GCAT cohort with their age-matching general population of Catalonia stratifying by age and sex, in order to assess their differences in sociodemographic, lifestyle and health-related factors, as we hypothesise that this cohort is affected by a healthy volunteer bias to some extent.

## Methods

### Design, setting and study population

This observational study compares the GCAT cohort with three administrative databases (SIDIAP, IDESCAT and INE, see below) representative of the general population of Catalonia, a Mediterranean region in North-east of Spain, with 7 675 217 inhabitants in 2019 according to the official population figures of the Municipal Register [9].

**GCAT cohort**. Is a population-based cohort of 20 000 participants from Catalonia, with active follow-up, who enrolled voluntarily in the study (2014-2018) with the unique restriction of being between 40 and 65 years old. Participants were recruited trough the national wide network of the Blood and Tissue Bank of Catalonia, but no restricted to blood donors. Specifically, informed participants agreed to participate in the study, provided informed consent and allowed access to the EHR from the public healthcare system for passive follow-up and to be contacted regularly to collect follow-up information on lifestyle and additional information. Participants can opt-out or withdraw their consent for specific areas of research. See cohort protocol description in Obón-Santacana et al 2018 [4]. The GCAT study was approved by the Germans Trias I Pujol University Hospital Ethical Committee (PI-13-020). **SIDIAP database**. The Information System for Performing Primary Care Research (SIDIAP) database collects pseudo-anonymized EHRs from 328 primary care centres in Catalonia managed by the Catalan Health Institute since 2005, and it currently has data on almost 80% of the Catalan population and is a reliable representation of the region in terms of age, sex and geographic distribution [10]. **IDESCAT**. Statistical Institute of Catalonia, since 1989, they provide official statistics of Catalonia, including social, demographic and economic data [9]. **INE**. National Statistics Institute, since 1945, provides official statistics of Spain, mainly on sociodemographic and economic characteristics [11].

### Inclusion and exclusion criteria

The participants included in the analysis met the following inclusion criteria: alive and living in Catalonia on the GCAT recruitment start date (2014), born between 1951 and 1970 to maximise the overlap between databases, and linked to EHR. Individuals detected to be duplicated, that opted out, or with discordant sex or birthdate were excluded. Finally, a total number of 13 434 GCAT and 1 625 075 SIDIAP participants were included in the comparison study.

### Domains and variables

Data collected included variables in fourth main domains (i) sociodemographic: sex, age, deprivation index [12, 13], educational attainment, civil status; (ii) lifestyle habits: smoking habit and alcohol consumption, (iii) health-related factors: mortality, primary care diagnosis, number of primary care visits; and (iv) medication dispensed in pharmacies: type of medication and quantity. Longitudinal comparisons were made until the end of 2019, or in the case of those variables measured at one-timepoint, the closest available register for the general population was obtained. Detailed information about all variables can be found in **Table S1**.

For this study, we grouped all primary care diagnosis into three-digits ICD-10 codes [14] and retained 20 diagnosis groups according to their chronicity, prevalence and/or relevance in terms of severity; as well as the 20 more prevalent types of cancer (see **Table S1**). For the medication, we selected all medication dispensed at the pharmacy grouped by second level ATC code [15] with at least 5% of prevalence in the general population, a total of 37. The final list of selected diseases include: Type 2 diabetes mellitus (E11), Overweight and obesity (E66), Disorders of lipoprotein metabolism and other lipidemias (E78), Alcohol related disorders (F10), Nicotine dependence (F17), Major depressive disorder, single episode (F32), Other anxiety disorders (F41), Migraine (G43), Essential (primary) hypertension (I10), Angina pectoris (I20), Chronic ischemic heart disease (I25), Atrial fibrillation and flutter (I48), Heart failure (I50), Cerebral infarction (I63), Atherosclerosis (I70), Vasomotor and allergic rhinitis (J30), Other chronic obstructive pulmonary disease (J44), Asthma (J45), Psoriasis (L40), Osteoporosis without current pathological fracture (M81).

### Statistical analysis

Statistical differences were computed with *Fisher’s exact test* or *t-test*. Individuals with missing data in any variable were excluded for that particular comparison. To calculate relative mortality and incidence disease rates by age, we corrected by the total number of individuals within the age range or older for each sex. Additionally, for mortality rate estimates, in those GCAT individuals that died during the recruitment we adjusted them for the proportion of individuals recruited. Lifetime prevalence, by sex, was computed as the number of individuals diagnosed at least once in a lifetime for a specific disease by the total number of individuals in the cohort.

### Weighting

We computed raked weights, an iterative proportional fitting procedure that adjusts one variable at a time [16, 17] for the GCAT cohort to be representative for the whole population. Calculation of weights was performed in R [18] using the anesrake package v0.80. Variables incorporated were age range, sex, deprivation index, education level, smoking habit and Charlson comorbidity index. This comorbidity index was calculated using the comorbidity R package v1.0.7 and then categorized according to their score (0, 1, 2+). Only individuals with no missing data in these variables were used in this calculation (n = 10 027).

## Results

### Sociodemographic characteristics

The GCAT cohort has 13 434 individuals fitting the inclusion criteria (69.3%), whilst SIDIAP has 1 625 075 (20.2%). GCAT participants have a significant higher proportion of female and are significantly younger (**Figure 1a, Table S2**).

**Figure 1.**
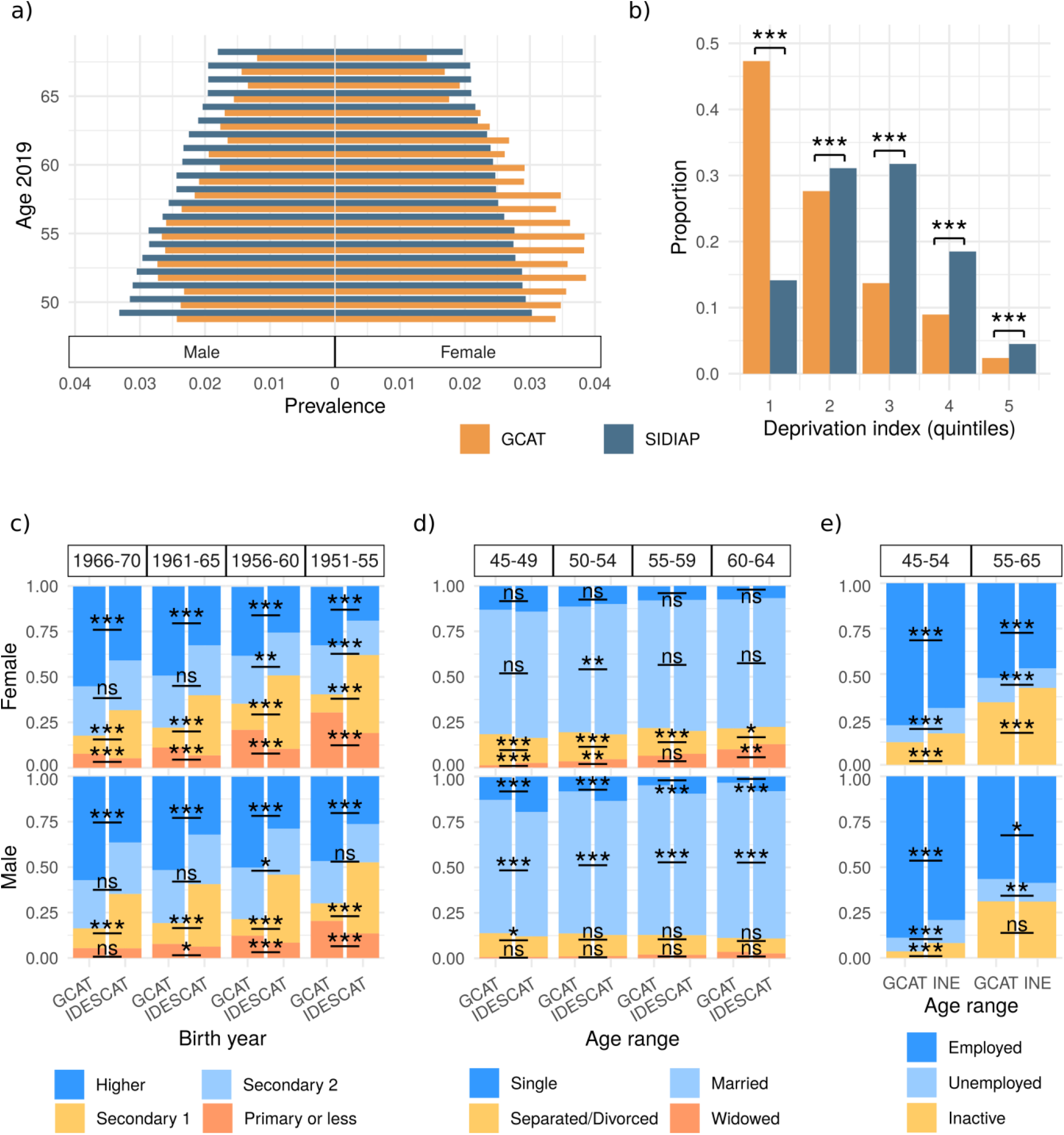
Comparison of sociodemographic characteristics between GCAT cohort and the general Catalan population. (**a**) Population pyramid comparison. (**b**) Deprivation index. In the x-axis quintiles of the deprivation index are represented, being 1 the inferior quintile (least deprived areas) and 5 the superior one (most deprived areas). In the y-axis the proportion of individuals is represented. (**c**) Education level in female (top) and male (bottom). In the x-axis the different sources and birth year groups are represented, and in the y-axis the proportion of individuals for each education level. (**d**) Civil status in female (top) and male (bottom). In the x-axis the different sources and age groups are represented and in the y-axis the proportion of individuals for each civil status. (**e**) Employment status in female (top) and male (bottom). In the x-axis the different sources and age groups are represented, and in the y-axis the proportion of individuals for each civil status.

GCAT participants live in less deprived areas than the general population (**Figure 1b, Table S2**), with a markedly higher proportion of individuals living in areas in the inferior quintile of deprivation index (less deprived).

GCAT participants have a higher proportion of individuals with higher education level than the general population for all sexes and age groups, although the proportion of individuals with primary education or less is also higher in GCAT participants, except for younger men where no differences are observed (**Figure 1c, Table S2**).

GCAT females are more divorced and less widowed than the general population. On the other hand, GCAT males have a higher proportion of married individuals and are less single than the general population (**Figure 1d, Table S2**).

Employment rates of GCAT participants are higher than the general population, with a lower proportion of labour inactivity than the general population. Unemployment rates are age-dependent, having a lower rate of unemployment than the general population at 45-54 age range and higher at 55-65 (**Figure 1e, Table S2**).

### Lifestyle habits

Alcohol consumption among GCAT participants is higher than the observed in the general population (**Figure 2a, Table S3**). The proportions of high- and low-risk drinkers are both higher in GCAT, except for younger men, with less high-risk drinkers, and concordantly, the proportion of non-drinkers is lower than the general population. Regarding smoking habits, the proportion of current smokers is lower in GCAT, the proportion of ex-smokers is similar to the general population in men and higher in women, and the proportion of non-smokers is markedly higher in men and slightly higher in women (**Figure 2b, Table S3**).

**Figure 2.**
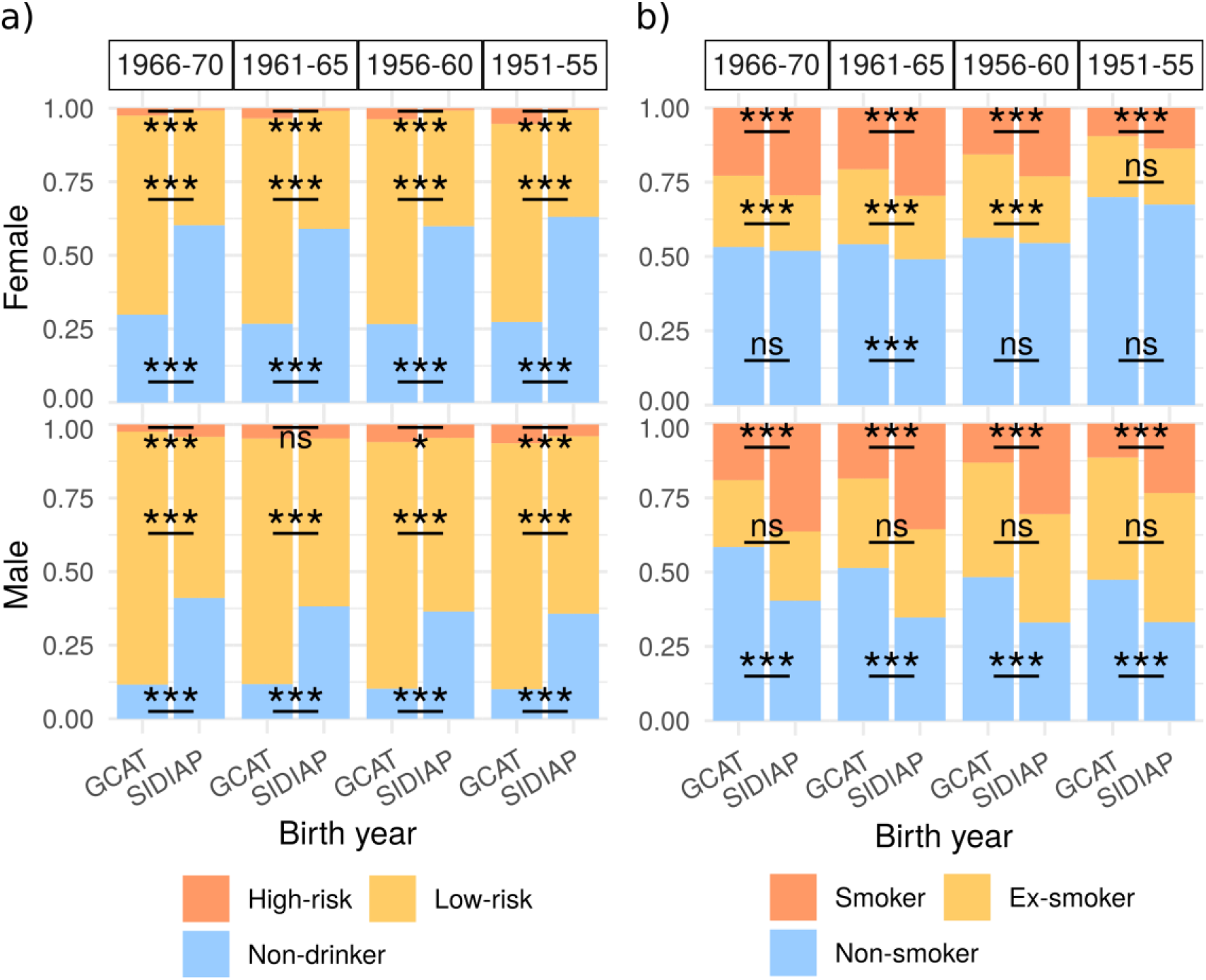
Comparison of lifestyle habits between GCAT Cohort and the general Catalan population. (**a**) Alcohol consumption in female (top) and male (bottom). In the x-axis the different sources and birth year groups are represented, and in the y-axis the proportion of individuals. (**b**) Smoking status in female (top) and male (bottom). In the x-axis the different sources and birth year groups are represented, and in the y-axis the proportion of individuals.

### Health-related factors

Mortality is lower in the GCAT cohort than in the general population for all sexes and age ranges (**Figure 3a, Table S4**).

**Figure 3.**
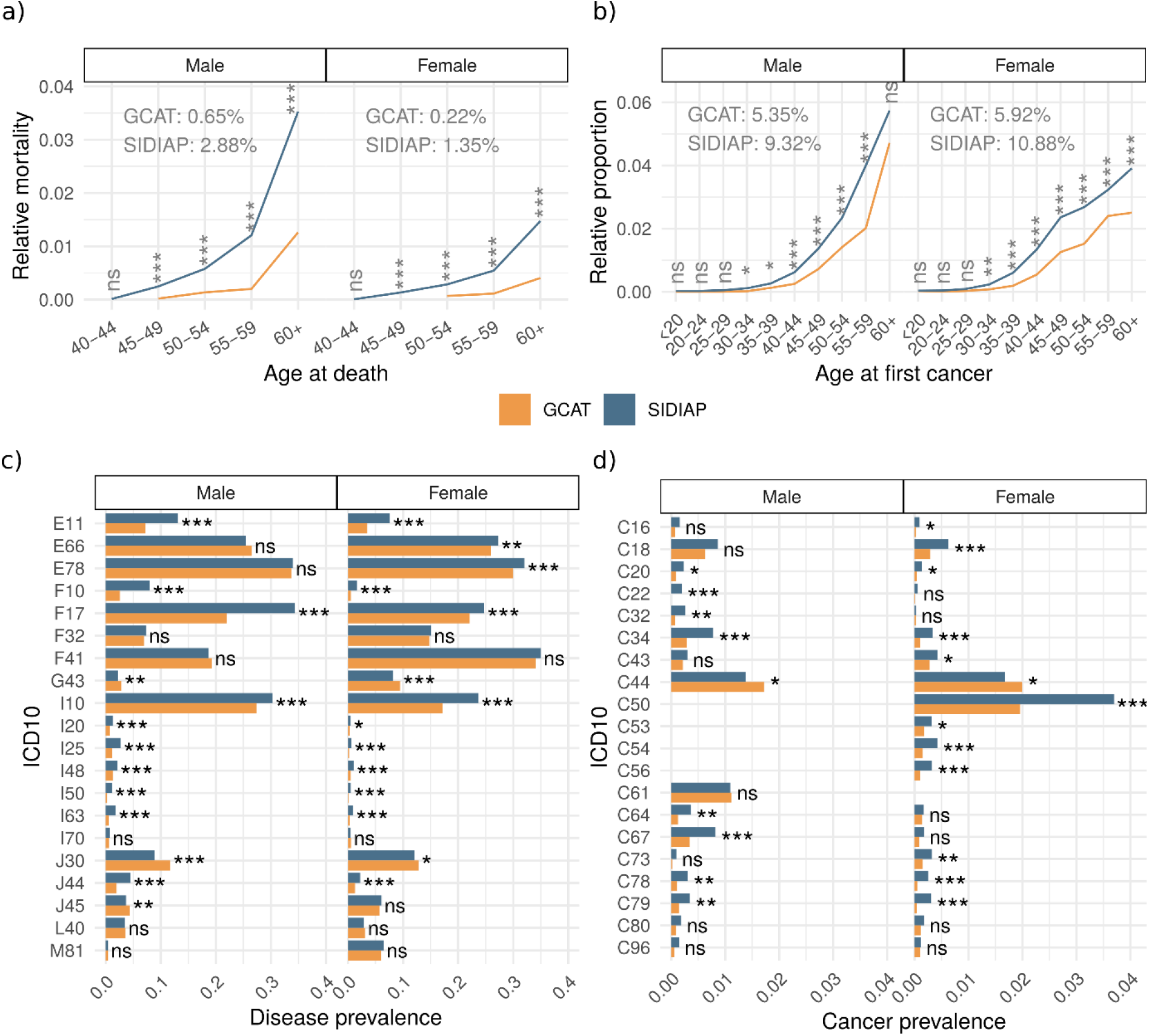
Comparison of health-related factors between the GCAT Cohort and the general Catalan population by sex and source. (**a**) Relative mortality by sex and source. In the x-axis the age range at death and in the y-axis the relative mortality during all the follow-up period. (**b**) Age at first cancer, including all malignant cancer types (ICD-10 codes C00-C99). In the x-axis the age range at first cancer diagnosis and in the y-axis the relative incidence. Overall cancer prevalence for each sex and source is included in the plot. (**c**) Disease prevalence. In the y-axis, the selected diseases (ICD-10 code) and in the x-axis the lifetime prevalence of each disease. (**d**) Cancer prevalence. In the y-axis, the different cancers (ICD-10 code) and in the x-axis the lifetime prevalence of each one.

When comparing the lifetime prevalence of the selected diseases (**Figure 3c, Table S4**), we can see differences in 15 out of 20 diseases between GCAT and the general population. GCAT individuals have lower prevalence of type 2 diabetes (E11), overweight and obesity (E66) in women, disorders of lipid metabolism (E78) in women, alcohol related disorders (F10), nicotine dependence (F17), essential hypertension (I10), angina pectoris (I20), chronic ischemic heart disease (I25), atrial fibrillation and flutter (I48), heart failure (I50), cerebral infarction (I63), COPD (J44). On the other hand, GCAT participants have higher prevalence of migraine (G43), vasomotor and allergic rhinitis (J30), and asthma (J45) in men.

In the case of cancer, the overall lifetime prevalence in GCAT is lower (female: P = 1.92e-52, OR = 0.52; male: P = 2.28e-28, OR = 0.55). Cancer incidence is lower in GCAT individuals older than 30, except for men aged 60+, where this difference is not significant (**Figure 3b, Table S4**). Regarding specific cancer types, we can highlight the differences in liver cancer (C22) in men, with no cases in GCAT, and the markedly lower prevalence in secondary neoplasms (C78, C79), bronchus and lung cancer (C34), ovarian (C56), uterus (C54), female breast cancer (C50), bladder cancer (C67) in men and colon cancer (C18) in women. On the other hand, non-melanoma skin cancer (C44) was the only cancer with a higher prevalence in GCAT (**Figure 3d, Table S4**).

The analysis of health services utilization was compared through diagnosis-associated primary care visits. GCAT participants of the younger age ranges have more primary care visits than the general population, while older individuals have the same number of visits than the general population (**Figure S1, Table S4**).

### Medication dispensed at the pharmacy

We compared all groups of medication (grouped by second level ATC code) with at least 5% of prevalence of use among the general population. We found that less GCAT individuals use medication for cardiovascular system (C codes), drugs used in diabetes (A10) and antidiarrheals, intestinal anti-inflammatory/anti-infective agents (A07) compared to the general population. On the other hand, there are more GCAT individuals using drugs for acid related disorders (A02), drugs for functional gastrointestinal disorders (A03) in men, vitamins (A11) and mineral supplements (A12) in women, drugs for blood and blood forming organs (B codes), dermatologicals (D codes), genitourinary system and sex hormones (G codes), corticosteroids for systemic use (H02), antibacterials for systemic use (J01), anti-inflammatory and antirheumatic products (M01), muscle relaxants (M03), drugs for nervous system (N codes), nasal preparations (R01), cough and cold preparations (R05) in men, antihistamines for systemic use (R06), ophthalomologicals (S01) and otologicals (S02) (**Figure 4a, Table S5**).

**Figure 4.**
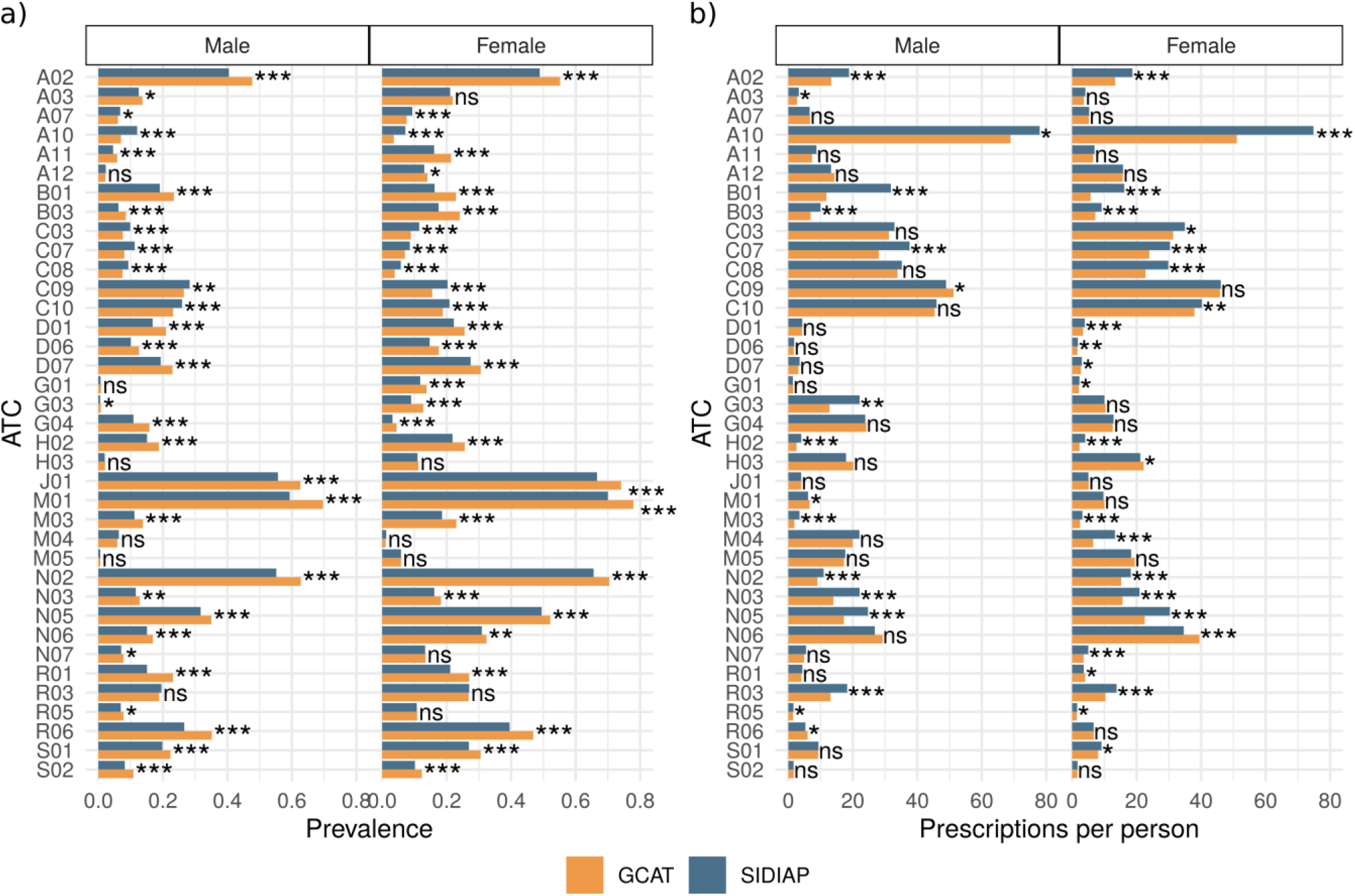
Comparison of medication use between GCAT Cohort and general Catalan population. (a) Drug use prevalence by cohort and sex. In the y-axis, the different groups of ATC codes and in the x-axis the prevalence of use for each group of drugs. (b) Number of mean prescriptions per person. In the y-axis, the different groups of ATC codes and in the x-axis the mean of prescriptions per person.

When comparing number of prescriptions per person taking that medication, GCAT individuals use less quantity of drugs than the general population. GCAT women use more psychoanaleptics (N06) and slightly more thyroid therapy (H03) and nasal preparations (R01), while GCAT men use slightly more agents acting on the reninangiotensin system (C09), anti-inflammatory and anti-rheumatic products (M01) and antihistamines for systemic use (R06). In all other medication groups, GCAT individuals use either less or similar quantity than the general population (**Figure 4b, Table S5**).

### Weighting

After applying raked weights, we can see that GCAT profile matches the one of the general population on those variables used to produce the weighting (**Table S6**). In the case of lifetime disease prevalence of the 20 selected diseases, we computed them before and after weighting (**Table S7**) and observed that the prevalence estimate improves in 16 of them, making it more similar to that of the general population than it was before weighting (**Figure 5**). In the four diagnosis groups with no improvement (Migraine, Atrial fibrillation and flutter, Atherosclerosis, and Osteoporosis), the difference between the general population and the estimated prevalence was under 1% in all of the cases.

**Figure 5.**
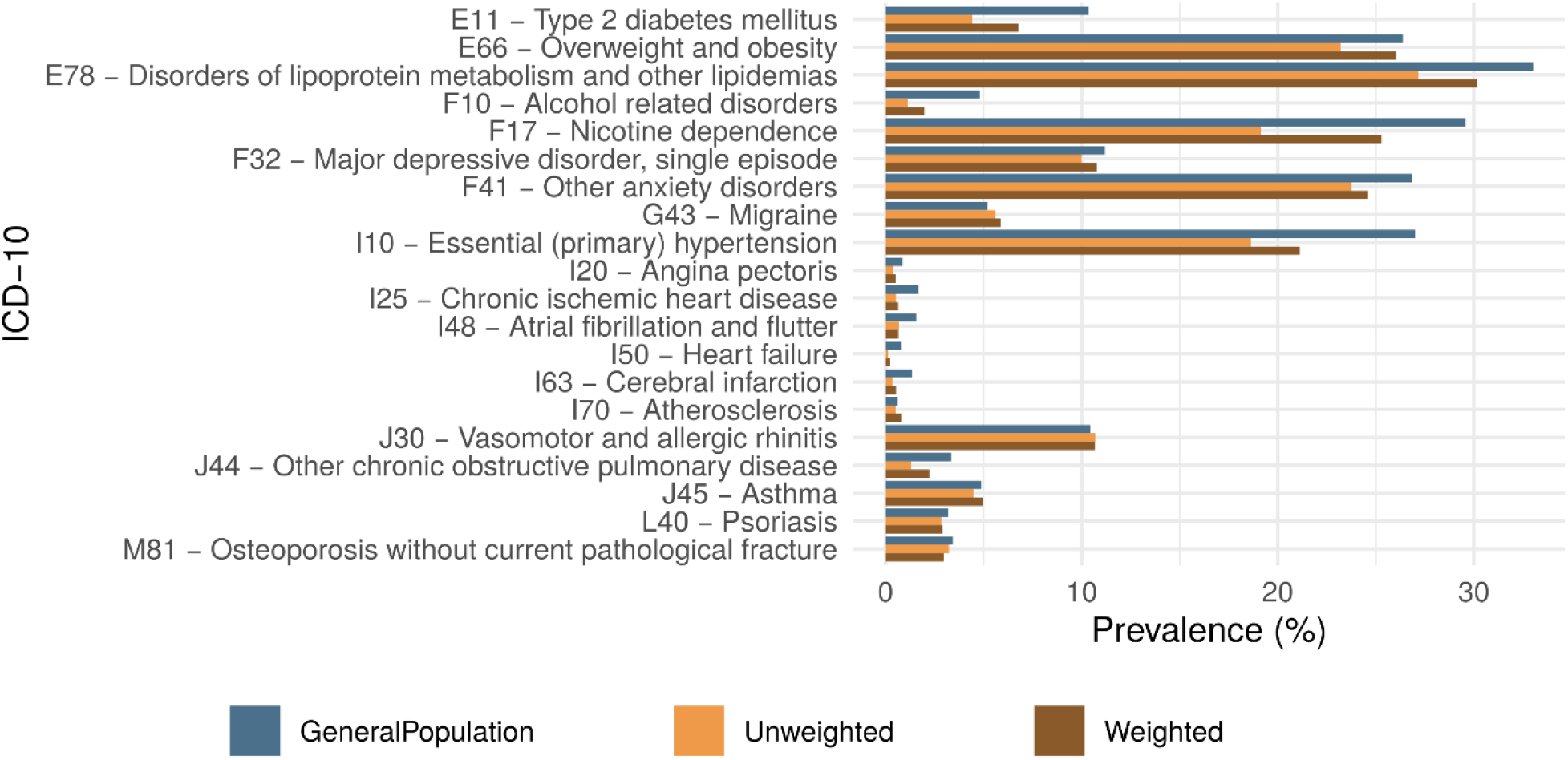
Bar plot of the lifetime disease prevalence between GCAT and the general Catalan population, before (yellow) and after weighting (brown) in a selection of diseases. In the x-axis the disease prevalence and in the y-axis the different diseases (ICD-10 code and description).

## Discussion

In this study, we have compared the GCAT cohort with the general population of Catalonia to assess its representativeness as a population-based Catalan cohort. We did this, determining statistical differences in a set of multiple variables across sociodemographic, lifestyle, disease and medication domains. As the description of the general Catalan population, we used the administrative SIDIAP database, encompassing 8 million users from primary healthcare attendance services, distributed across the country, and then compared with individuals of the GCAT cohort with available linked EHR data. In addition, we used IDESCAT and INE databases, statistical institutes from Catalonia and Spain respectively, to compare some sociodemographic characteristics that were not available in SIDIAP.

We found differences in all the compared domains and produced weighted scores to extrapolate results to the general population based on biased participation. In brief, it has been observed that in the GCAT cohort, volunteers have a higher education level and live in less deprived areas compared to the general population. Women and younger individuals seem to be overrepresented compared to the general population. The mortality in the cohort is markedly lower and, in concordance, a lower prevalence of frequent non communicable diseases, as cancer, endocrine and cardiovascular diseases is observed among volunteers from the cohort. Regarding toxic habits, smoking prevalence is lower than in the general population but alcohol users are overrepresented. Furthermore, when looking at medication, among GCAT participants, drugs are less used than in the general population, but we observe a wider variety in drug use.

The observed gender, socioeconomic and healthy bias have been already observed in other population-based cohorts [19-21]. Gender bias, with a systematic overrepresentation of female participants, has been explained by the reverse gender gap in volunteer activities, described to be associated with cultural gender roles and inequality [22]. Regarding age bias, there is heterogenicity in this type of studies, in our case, we found an overrepresentation of younger individuals in comparison to the general population, but given that they were mainly blood donors (97%), it was expected to have a slightly higher number of younger individuals, as individuals between 45-54 represent the 24% of blood donors and considerably declines at older age groups (18.15% in 55-59 age groups and 10.52% in 60-69 age groups, corrected by the number of years in the period) [23]. Despite this drop in older individuals blood donations, the decrease in GCAT participation was not that marked for this age range, finding an overrepresentation of older women compared to blood donors, probably due to less time constraints. A higher socioeconomic status was observed, correlated with a higher education level and being employed, that although it is usually associated more time constraints, they show a higher interest in voluntarism as well as in science, being aware of the importance of these type of studies [24]. Regarding marital status, the increase in divorce rates in female can be explained by an increased economic independence [25], decrease in widowed female could be due to healthy partners, in concordance with participants itself, and increase in male marriages can be explained by the already described relationship between married people and good health [26].

The healthy bias, previously described as healthy volunteer effect, including less mortality and higher health status [6, 7], is partly associated to an increased health-consciousness and preventive behaviour of individuals with a higher socioeconomic status, leading to a higher proportion of healthy behaviours, as observed lower prevalence of current smokers, a higher diversity of drugs used in a lower quantity, and a higher number of healthcare visits, although this does not translate into a higher number of chronic diagnoses. Deprivation index, as a proxy of socioeconomic status has been associated with better health related parameters in a way that living in high deprived areas increase the likelihood of having more than one long-term condition at the same time, and on average people in the most deprived quintile of the population develop multiple long-term conditions 10 years earlier than those in the least deprived quintile [27].

In this scheme of healthier risk habits, we observed that alcohol consumption was higher in GCAT participants compared to those of the general population, however when comparing alcohol related disorders diagnosis (ICD-10 code F10) we observed a lower prevalence in the GCAT cohort. This could be explained by the diversity of sources: a quantitative estimation from a detailed alcohol consumption (based on a validated questionnaire) in the GCAT cohort versus an EHR-based qualitative estimation by the physician and/or the nurse. In addition, the observed differences in alcohol consumption can be attributed to the mode effect, as people tend to report more desirable answers in the presence of an interviewer (i.e., the physician) [28]. A similar trend was observed in smoking habit, were both sources are available (**Figure S2**). This result outlines the urgent need for a better register of the risky habits during the recurrent healthcare visits. If we want to develop personalized support tools that consider personal risk behaviours, real world data solutions should be incorporated in the current practice.

Regarding health status, we observed an overall lower prevalence of chronic diseases in the GCAT cohort. We observed a lower prevalence in alcohol related diseases and nicotine dependence, as previously discussed, and a lower prevalence in endocrine and cardiovascular diseases, two related groups of diseases, being endocrine diseases a risk factor for cardiovascular diseases [29], strongly affected by lifestyle habits, especially diet, physical activity, poor sleep and smoking habit [30]; and socioeconomic status, due to increased levels of behavioural and psychosocial risk, and they usually come from low socioeconomic status families too, having a poor health during childhood [31, 32].

A similar healthy trend is observed in cancer prevalence, this is expected due to healthier lifestyle habits and more health-consciousness of the GCAT participants, as observed in other general-population studies [21], but keeping in mind the high proportion of blood donors in the cohort, it can also be affected by the impossibility to donate blood during or after a cancer, with few exceptions [33], affecting the number of prevalent cases at recruitment. Interestingly, non-melanoma skin cancer is more prevalent in GCAT individuals. An association with higher socioeconomic status was already observed, in particular in basal cell carcinoma, a type of non-melanoma skin cancer, associated with intermittent sun exposure and sunburn, probably due to more leisure time, leading to outdoor activities and holidays in the sun, as well as for the socially favoured perception of tanning [34]. This exposure-disease association was previously observed in the GCAT cohort [35].

Overall, this study has important strengths, as is the use of EHR data for SIDIAP and GCAT coming from the same source and being totally comparable in a large number of variables, or the generation of weights to make results generalisable to the entire population. In addition, gender and age-range stratification allows accounting for gender and age-related differences in biology of disease and risky and social behaviour. However, is important to note that there are important limitations, as the use of only primary care registries, and the incompleteness of registries before 2010. Using this data, do not allow a precise estimation of the age at first diagnosis in early onset diseases (usually diagnosed before 2010), but the approach is confident for late-onset diseases comparison. It is important to note that there is some information that is not available for the general population, or it is aggregated, and it can’t be properly compared due to incompatible age ranges or lack of sex stratification. Sample overlap could affect the use of Fisher’s exact test and t-test, given that GCAT individuals are mostly included in the SIDIAP database. However, this overlap is under 1%, having a meaningless impact on test performance [36]. Although it is a limitation for some types of analysis, it should not be affecting exposure-disease associations, as long as the different levels of exposure are properly represented in the cohort [37].

In conclusion, the integration of multidomain weights, which encompassed a wide range of factors including sociodemographic traits, lifestyle choices and health-related indicators, has successfully addressed the inherent bias observed in disease prevalent estimates within the GCAT cohort. This strategic correction has yielded an improved alignment of the cohort’s characteristics with those of the general Catalan population, thereby enhancing its representativeness and overall applicability.

## Declarations

### Ethics approval

The GCAT study was approved by the Germans Trias I Pujol University Hospital Ethical Committee (CEIC-HGTP) on April 26^th^, 2013 (PI-13-020). This study with SIDIAP Data was approved by the Scientific and Ethical Committees on December 18^th^, 2019 (19/518-P).

### Data availability

Original data is available upon request to the authors (correspondence author). All the data generated during this study (results) is available as supplementary data.

## Acknowledgements

This study makes use of data generated by the GCAT Genomes for Life, a cohort study of the Genomes of Catalonia, Fundació IGTP. IGTP is part of the CERCA Program / Generalitat de Catalunya. This study was carried out using anonymized data provided by the Catalan Agency for Quality and Health Assessment, within the framework of the PADRIS Program. The authors of this study would like to acknowledge all GCAT project investigators who contributed to the generation of the GCAT data. A full list of the investigators is available from www.genomesforlife.com/. We thank the Blood and Tissue Bank from Catalonia (BST) and all the GCAT volunteers that participated in the study. We gratefully acknowledge Beatriz Cortés (2019-2022) and Anna Carreras (2012-2022), former GCAT workers for their contributions to this study.

