## Supplemental Figures for "Disease prevalence, health-related and socio-demographic factors in the GCAT cohort. A comparison with the general population of Catalonia"

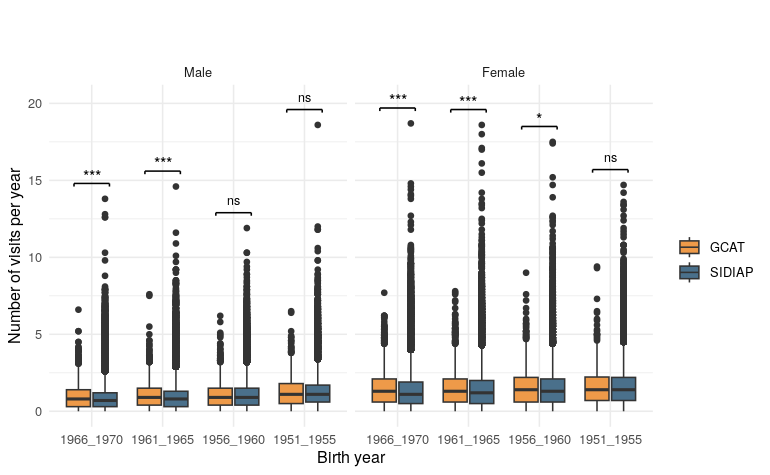
**Fig. S1** Boxplot of the number of primary care visits per year by sex and source. In the x-axis, the birth year, in the y-axis the number of visits per year.

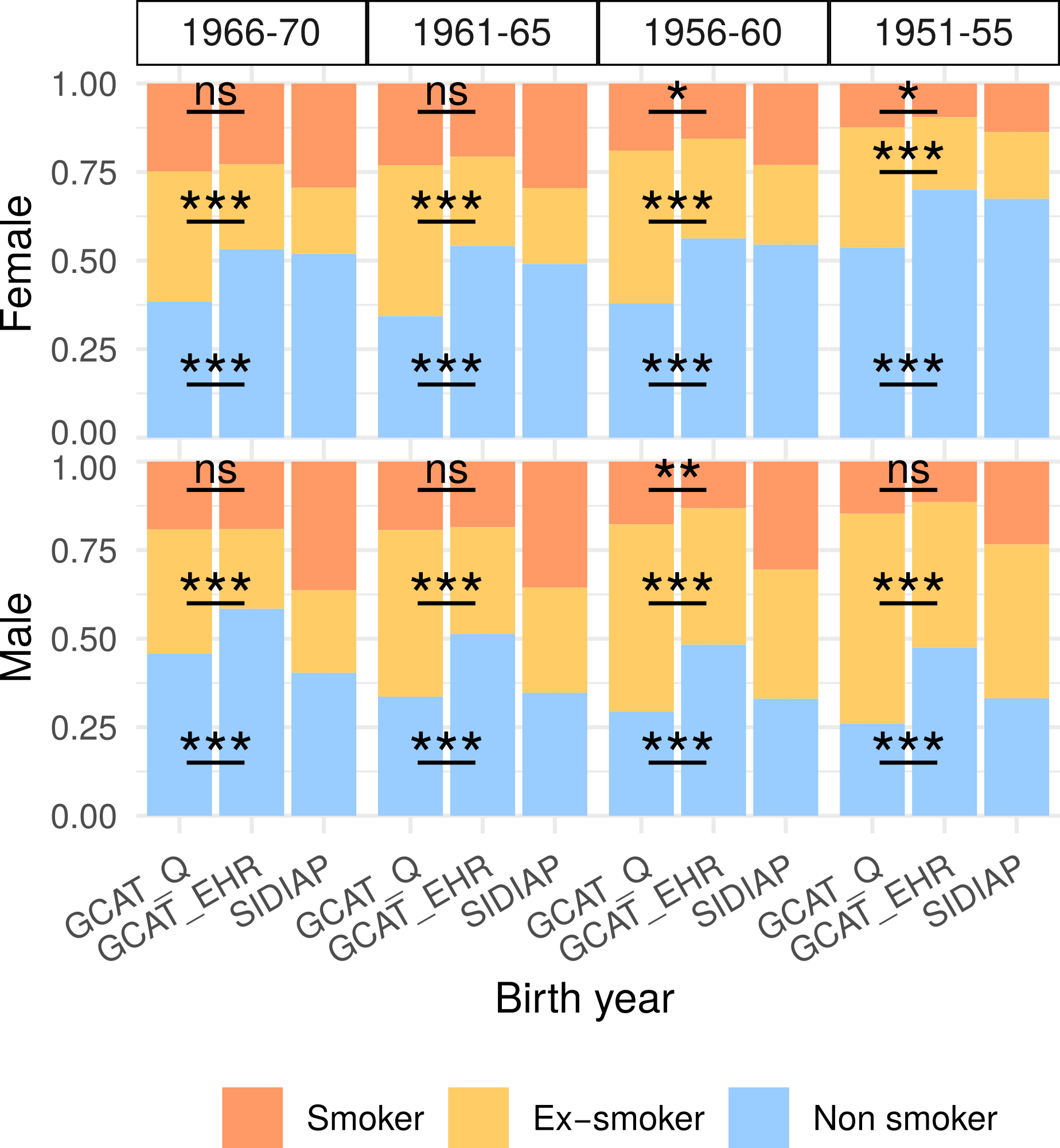

**Fig. S2** Stacked bar plot for smoking status in female (top) and male (bottom). In the x-axis the different sources and birth year groups are represented, in the y-axis the proportion of smoking status in the questionnaire based (GCAT_Q) and the EHR one (GCAT_EHR). We included SIDIAP, with the same EHR source, to provide a reference.
